# Impact of Cardiopulmonary Resuscitation Duration Prior to Extracorporeal Support on Mortality After Surgery for Acute Type A Aortic Dissection with Cardiopulmonary Arrest

**DOI:** 10.64898/2026.02.18.26346593

**Authors:** Soichiro Kageyama, Takeki Ohashi, Masahiko Kuinose, Tomoki Yamatsuji, Taiki Kojima

## Abstract

**Background:** Acute type A aortic dissection (AAAD) complicated by cardiopulmonary arrest is characterized by high mortality rates, rendering the selection of surgical candidates a subject of intense debate. Despite the necessity for cardiopulmonary resuscitation (CPR) prior to the completion of a definitive intervention, the prognostic impact of CPR duration on postoperative survival and neurological outcomes remains insufficiently elucidated. This study sought to evaluate the association between pre- and intra-operative CPR duration and the incidence of early mortality and central nervous system (CNS) complications in patients undergoing emergent surgical repair for AAAD.

**Methods:** This retrospective, cohort study was conducted at two tertiary community hospitals in Japan. All the patients who underwent emergency surgery for AAAD between January 2014 and December 2024 were enrolled. A multilevel Cox proportional hazards model, with each patient as level 1 and institutions as level 2, was used to evaluate the association between pre-or intra-operative CPR events and early postoperative mortality and CNS complications.

**Results:** Of the 880 patients enrolled, 785 (89.2%), 13 (1.5%), and 82 (9.3%) were without CPR, with CPR <15 min, and with CPR ≥15 min, respectively. Among them, death within 30 days post-surgery occurred in 76/785 (9.7%), 3/13 (23.1%), and 47/82 (57.3%), respectively. CNS complications within 30 days post-surgery occurred in 141/785 (18.0%), 5/13 (38.5%), and 38/82 (46.3%) without CPR, CPR <15 min, and ≥15 min, respectively. In multivariable analysis, CPR lasting ≥15 min was associated with mortality within 30 days post-surgery (adjusted hazard ratio, 7.66; 95% confidence interval [CI], 3.56–16.5; P<0.001). Both CPR <15 min and ≥15 min were associated with an increase in the sub-hazard ratio of CNS complications within 30 days post-surgery (adjusted sub-hazard ratios, 4.49; 95% CI, 3.92–5.11; P<0.001, and 3.62; 95% CI, 2.73–4.81; P<0.001, respectively).

**Conclusion:** A preoperative CPR duration of ≥15 min prior to the initiation of cardiopulmonary bypass or extracorporeal membrane oxygenation was associated with a substantial escalation in 30-day mortality compared with patients without CPR. These findings suggest that CPR duration might serve as a pivotal prognostic indicator, necessitating careful consideration for surgical indication in patients with AAAD complicated by CPR.

**CLINICAL PERSPECTIVE:** *What is new?:*

- Pre- or intra-operative cardiopulmonary resuscitation lasting ≥15 min in patients with acute type A dissection is associated with a nearly seven-fold increase in 30-day postoperative mortality.
- Both short (<15 min) and prolonged (≥15 min) cardiopulmonary resuscitation are associated with a higher risk of early postoperative complications in the central nervous system.

*What are the clinical implications?:*

- Patients with acute type A dissection who require pre- or intra-operative cardiopulmonary resuscitation ≥15 min should undergo careful multidisciplinary evaluation, as the risk of early mortality is substantially elevated.
- Even brief cardiopulmonary resuscitation is associated with increased neurological complications, highlighting the need for early neurological monitoring and supportive care postoperatively.

## 1. INTRODUCTION

Acute type A aortic dissection (AAAD) is a medical emergency with high mortality and high complication rates, especially in cases with preoperative cardiopulmonary arrest (CPA).^1–5^ For surgically treated patients with AAAD experiencing pre- or intra-operative CPA, the indications for emergency surgery remain controversial because of the limited evidence regarding postoperative mortality and neurological outcomes.

AAAD is often challenging to diagnose in patients presenting to the emergency room with CPA, and some of these patients may die before diagnosis and thus are not captured in the AAAD registry. Therefore, the actual preoperative mortality rate or appropriateness of treatment may be underreported. In addition, when the AAAD is diagnosed, emergency open-surgery is offered as the only treatment option for potential survival,^6^ and the profound multiple organ damage due to preoperative CPA raises concern that surgical treatment may be futile in specific individuals. Despite the clinical urgency, no consensus guidelines currently define surgical indications, exclusion criteria, or acceptable thresholds of preoperative resuscitation in AAAD complicated by CPA.

Extremely high mortality rates were observed in cases where return of spontaneous circulation (ROSC) was not achieved preoperatively or where cardiopulmonary resuscitation (CPR) duration exceeded 15 min^6–9^; however, these findings are limited to small-scale retrospective studies. Prior studies have not established reliable prognostic markers, and critical problems, such as the maximum duration of preoperative CPR or exclusion criteria for emergency surgery, remain unresolved. In addition, central nervous system (CNS) outcomes are unknown. CNS prognosis is equally important in deciding surgical candidacy, with significant implications on quality of life, long-term functional outcomes, and ethical considerations. Thus, cardiovascular surgeons currently make time-critical decisions based on minimal evidence to select candidates who may still benefit from emergency open surgery.

Thus, in this study, we aimed to evaluate the association between CPR administration until initiation of cardiopulmonary bypass (CPB) or extracorporeal membrane oxygenation (ECMO) and early postoperative mortality and neurological outcomes in patients with AAAD. We hypothesized that a longer duration of CPR would be associated with increased early postoperative mortality and CNS complications.

## 2. MATERIALS AND METHODS

### 2.1 Study Design, Setting, and Population

This multicenter, retrospective, cohort study was conducted in two tertiary-care community hospitals in Japan between January 2014 and December 2024. This study was approved by the centralized Institutional Review Board. The local Ethics Committee of the Kawasaki Medical School Hospital (approval number: 6796-01) approved this study on June 27, 2025. This study included all adult patients aged ≥18 years who underwent emergency surgery for AAAD. This retrospective observational study was exempt from obtaining written patient consent because it utilized existing patient information. This cohort study followed the Strengthening the Reporting of Observational Studies in Epidemiology (STROBE) guidelines.^10^

### 2.2 Data Collection

Data on patients’ underlying characteristics and surgical procedures were extracted from the electronic medical records at the participating institutions. The primary investigator (SK) retrospectively extracted and recorded the data for analysis, using predefined definitions. This study obtained the following information for the analysis: date of surgery, last date of a clinic visit or hospital admission (death during the admission), patient demographics (age, sex, history of hypertension, diabetes mellitus, dyslipidemia, renal dysfunction, hemodialysis, aortic aneurysm, Marfan’s syndrome, aortic dissection, and cardiac surgery), preoperative complications (unstable hemodynamics, major organ ischemia), entry and type of aortic dissection, preoperative patient status (cardiac function, presence of coagulopathy, and thrombocytopenia), surgical characteristics (location of aortic placement, additional procedures, duration of surgery and CPB, and presence of massive transfusion), postoperative complications (chest reopening, CNS disorder, infection, renal disorder, gastrointestinal disorder, multiple organ failure, and re-dissection and re-rupture of large vessels), and postoperative extracorporeal circulatory support (ECMO and left ventricular assisting device).

### 2.3 Definitions

All the study variables were defined as in our previous report.^4^ **“CPR time”** was defined as the time from the initiation of CPR until 1) ROSC was achieved; 2) ECMO was attached before surgery; or 3) CPB was started during surgery. **“CNS complications”** was a composite outcome, including focal neurological disorder, consciousness disorder, convulsion, and other CNS disorders (i.e., cerebral infarction, dysphagia, cognitive dysfunction, visual field deficit, subarachnoid hemorrhage, cerebral hemorrhage, recurrent nerve paralysis, and neurogenic bladder). **“Cardiac tamponade”** was defined as preoperative hemodynamic instability attributable to aortic rupture. In cases where other potential causes of shock, such as coronary ischemia or multiorgan failure, were concurrently present, the final diagnosis of cardiac tamponade was determined based on intraoperative findings, including the sudden release of hemorrhagic pericardial effusion and associated hemodynamic changes following pericardiotomy and drainage. Other definitions of research terminologies are described in Supplemental Text 1.

### 2.4 Preoperative Diagnosis

The diagnosis of AAAD was established primarily through preoperative contrast-enhanced computed tomography (CT). In cases of hemodynamic instability, an expedited non-contrast CT scan was performed instead of a contrast-enhanced imaging. For patients who were transported emergently to the operating suite due to critical clinical status, the diagnosis of AAAD had been confirmed at the referring institutions using aortography combined with coronary angiography, supplemented with additional imaging modalities, including contrast-enhanced CT.

### 2.5 Indication for Surgery

#### 2.5.1 Surgical Policy and Perioperative Management

Surgical intervention was considered for all patients with AAAD, regardless of aortic diameter or false lumen status on preoperative imaging. The decision to proceed with surgery was based on an individualized assessment incorporating patient preferences, family input, and baseline functional status. Most patients with AAAD at our institution are referred from other hospitals. Upon receiving a referral, a cardiovascular surgeon from our hospital is dispatched to the referring facility via a physician-staffed emergency vehicle (doctor car).

The surgeon confirms the diagnosis, determines the indication for emergency surgery, and initiates the transfer. To minimize delays, the operating room is prepared concurrently during transport. Patients in unstable condition are transferred directly to the operating room upon arrival.

#### 2.5.2 Management of Cardiopulmonary Arrest

Indications for emergency surgery in patients with CPA are evaluated at several critical points: upon the surgeon’s arrival at the referring hospital, upon arrival at our facility, and the earliest stage of surgery with CPB initiated. If CPA occurs at the referring hospital, transport is generally canceled if CPR has exceeded 30 min by the time our surgeon arrives. However, if the CPR duration is less than 30 min, transport is initiated while continuing CPR.

For patients who remain in CPA upon arrival at our hospital, the decision to proceed with emergency surgery is made based on the duration of CPR and the family’s informed consent. Emergency open surgery is performed if the patient achieves ROSC before arrival or if the total CPR duration is under 30 min.

After transferring to the operating room, prompt thoracotomy and CPB initiation are performed, followed by a final decision regarding continuation of surgical intervention based on cardiac activity recovery, presence of organ dysfunction, and neurological assessment.

The initiation of veno-arterial ECMO as a bridge to surgery is determined by institutional factors, including operating room readiness, catheterization laboratory availability, and staffing levels. When chest compressions are ineffective due to massive pericardial tamponade, pericardial drainage may be performed at the referring hospital or in our emergency department. However, our priority is to avoid any delays in transport or the commencement of definitive open surgery.

#### 2.5.3 Surgical Indications

At the participating institutions, the surgical management of AAAD complicated by preoperative CPA follows a systematic, stepwise algorithmic approach. Patients requiring CPR >30 min upon arrival are generally deemed ineligible for surgical intervention. For those with a CPR duration <30 min, resuscitation is maintained during transfer to the operating suite, with ECMO being instituted as clinically indicated. Patients already under ECMO support are transferred directly for emergent surgery. Following immediate thoracotomy and the establishment of CPB, surgical futility is rigorously reassessed. In the absence of spontaneous cardiac activity despite the relief of tamponade and CPB initiation or in the event of a myocardial “stone heart,” the decision to proceed is contingent upon a multidisciplinary evaluation of preoperative organ dysfunction and neurological integrity, including the level of consciousness and pupillary light reflexes. Consequently, the definitive “point of no return” for surgical completion is established at the time of CPB initiation; therefore, this study defined a combination of pre- and intra-operative CPR duration until the initiation of CPB as the interval leading up to this critical juncture.

In principle, we employ a conventional tear-oriented approach to determine the extent of graft replacement.^4^ For patients who experienced preoperative CPA, we prioritize minimally invasive procedures to account for systemic instability, such as multi-organ failure or coagulopathy. Consequently, concomitant cardiac procedures for valvular disease or coronary artery disease are avoided unless the conditions are life-threatening.

### 2.6 Surgical Procedures

Tear-oriented surgery was performed at the two participating institutions. The intimal tear was excised, and the extent of aortic replacement was determined by its location. If no tear was identified in the ascending aorta or aortic arch, replacement was limited to the ascending aorta. Extended aortic replacement was performed if the dissection or rupture extended beyond the primary tear. The femoral artery was the preferred site for arterial cannulation; however, the axillary artery was selected via a side-graft if aneurysmal changes, intimal irregularities, or significant atheroma were present in the descending or abdominal aorta.

Venous drainage was achieved through the right atrium or the superior and inferior vena cava. The proximal aortic stump was prepared by obliterating the false lumen with fibrin sealant (Beriplast P Combi-Set; CSL Behring, Tokyo, Japan). The aortic wall was reinforced using Teflon felt strips placed in a “sandwich” configuration, secured with 4-0 polypropylene horizontal mattress sutures (Surgipro II; Covidien, Mansfield, MA). After completing a continuous 3-0 polypropylene suture, BioGlue Surgical Adhesive (CryoLife, Kennesaw, GA) was applied to the anastomotic sites to ensure hemostasis at the needle holes.

During CPB, the ascending aorta was clamped and incised. Cardioplegic solution was administered directly into the coronary ostia to achieve cardiac arrest, followed by trimming of the proximal aortic stump. Once the rectal temperature reached 28 °C, systemic circulatory arrest was initiated, the aorta was unclamped, and selective cerebral perfusion was started.

The distal aortic stump was then trimmed and anastomosed to the vascular prosthesis, after which systemic perfusion was resumed via the prosthetic side branch. In cases of total aortic arch replacement, the arch vessels were reconstructed individually. Finally, rewarming was initiated, proximal anastomosis was completed, aortic clamp was removed, and the patient was weaned from CPB.

### 2.7 Outcomes and Exposure

The primary outcome was death from any cause within 30 days of surgery. The secondary outcomes were death from any cause within 7 days of surgery and CNS complications within 30 days of surgery. The exposure was the administration of CPR, trichotomously categorized as 0, <15 min, or ≥15 min.

### 2.8 Statistical Analysis

For descriptive statistics, categorical variables are reported as numbers and percentages. Non-normally distributed continuous variables are reported as median [interquartile range (IQR)].

For bivariable analysis, the chi-squared and Fisher’s exact tests were applied to categorical variables, and analysis of variance (ANOVA) or the Kruskal–Wallis test was applied to continuous variables with normally or non-normally distributed data, respectively.

The exposure variable (CPR administration) was categorized into three levels (none, <15 min, and ≥15 min). Kaplan–Meier estimation was used to evaluate cumulative survival probabilities at 7 and 30 days after surgery across the three CPR performance categories. Multilevel Cox proportional hazards models were used, adjusting for potential confounders as fixed effects and the institution-level variance as random effects, with individual patients at level 1 and institutions at level 2.^11^ The potential confounders were selected based on clinical perspectives by board-certified cardiac surgeons and knowledge from literature,^12–20^ including age, sex, preoperative CNS complications, preoperative renal dysfunction, preoperative ischemic cardiac conditions/ tamponade, duration ≥30 min between attachment of preoperative ECMO and initiating CPB, DeBakey type Ⅱ dissection, and thrombosed false lumen. For secondary analysis, to evaluate the association between the occurrence of CNS complications within 30 days after surgery and CPR administration, the competing risk exists between death and the occurrence of CNS complications within 30 days after surgery. To address this competing risk, the Fine and Gray proportional subhazards regression analysis was applied,^21^ adjusting for the same potential confounders as fixed effects in the primary analysis and accounting for within-cluster correlation at each institution using standard errors.

The cohort who received CPR <15 min included only a small number of participants (n=13), which can make risk estimates in the regression models unstable owing to insufficient power. Thus, a sensitivity analysis excluding participants with CPR time <15 min was conducted.

ECMO attachment and ROSC provide physiologically different circulatory conditions that can affect the outcomes. Thus, we conducted a sensitivity analysis in the cohort, excluding participants who underwent ECMO attachment.

A complete case analysis was applied to conduct multivariable regression analyses, as there was a small number of cases (five cases) with missing values in the covariates that were incorporated into the regression analyses. Data were analyzed using STATA 18.0 (StataCorp, College Station, TX), with a two-sided P-value <0.05, as the criterion for statistical significance.

## 3. RESULTS

### 3.1 Study Population

Between January 2014 and December 2024, 880 patients underwent emergency surgery for AAAD at the two recruited hospitals. A total of 785/880 (89.2%) patients had no pre- or intra-operative CPR, 13/880 (1.5%) had <15 min of pre- or intra-operative CPR, and 82/880 (9.3%) had ≥15 min of pre- or intraoperative CPR.

Preoperative cardiac tamponade and ECMO attachment were present in 10/13 (76.9%) and 45/82 (54.9%) in the CPR <15 min and CPR ≥15 min groups, respectively; whereas they were absent in the patients without CPR events. Preoperative coronary and cerebral ischemia were found approximately 2–3 times in the two groups who received CPR compared with the group who did not receive CPR. Fewer patients with a thrombosed false lumen were identified in the group with ≥15 min of CPR events compared with the groups without CPR events or with CPR events <15 min (Table 1). Regarding surgical characteristics, the durations of CPB, circulatory arrest, and selective cerebral perfusion were approximately 1.2 to 1.5 times longer, and deep hypothermic cardiac arrest was performed approximately 2.5 times as often in the group with CPR ≥15 min compared with the group without CPR events (Table 2). Overall, the proportions of renal failure with dialysis, multiple organ failure, and extra circulation support are predominantly identified in the group with CPR ≥15 min compared with the group without CPR events (Table 3).

**Table 1.**
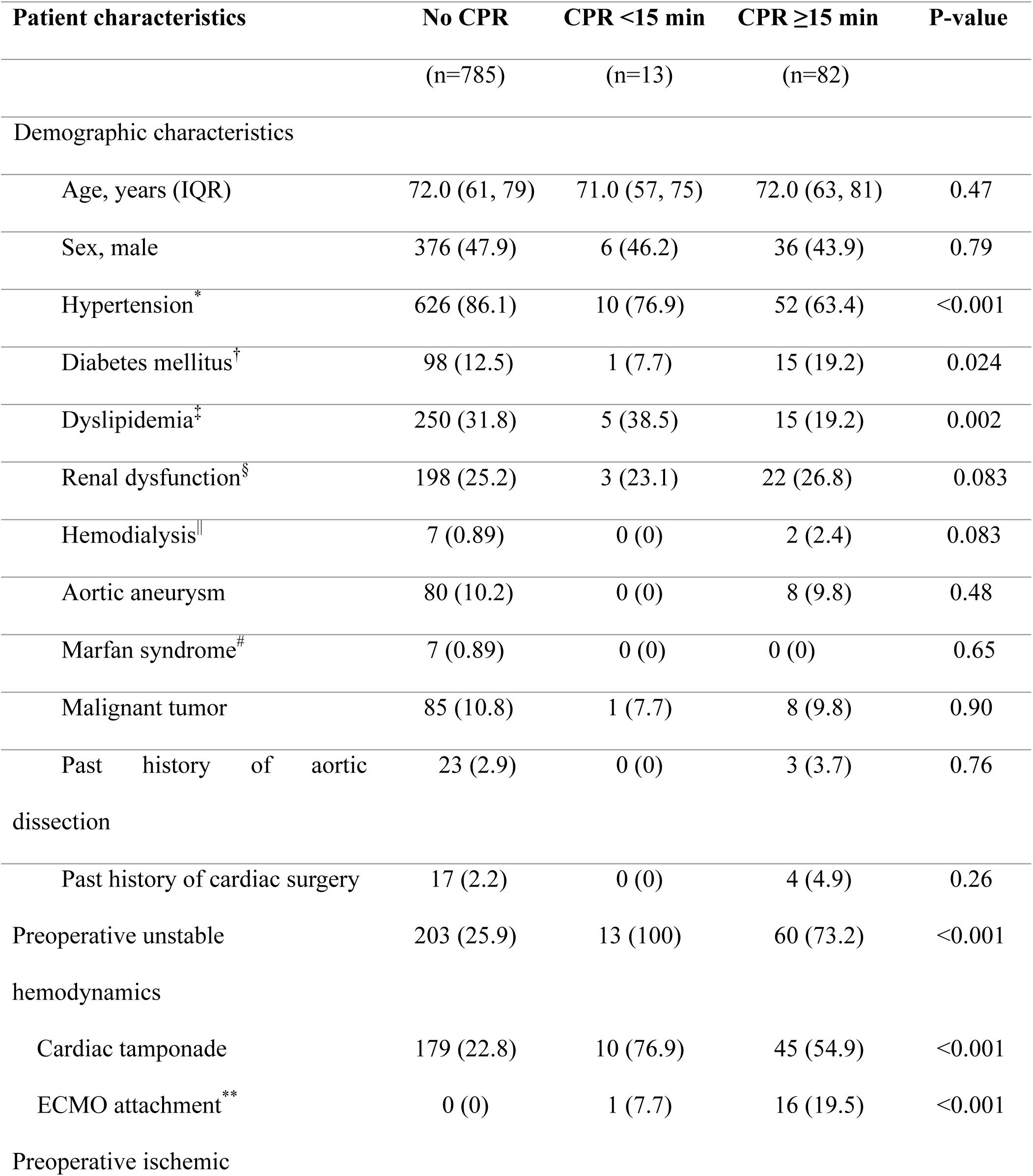
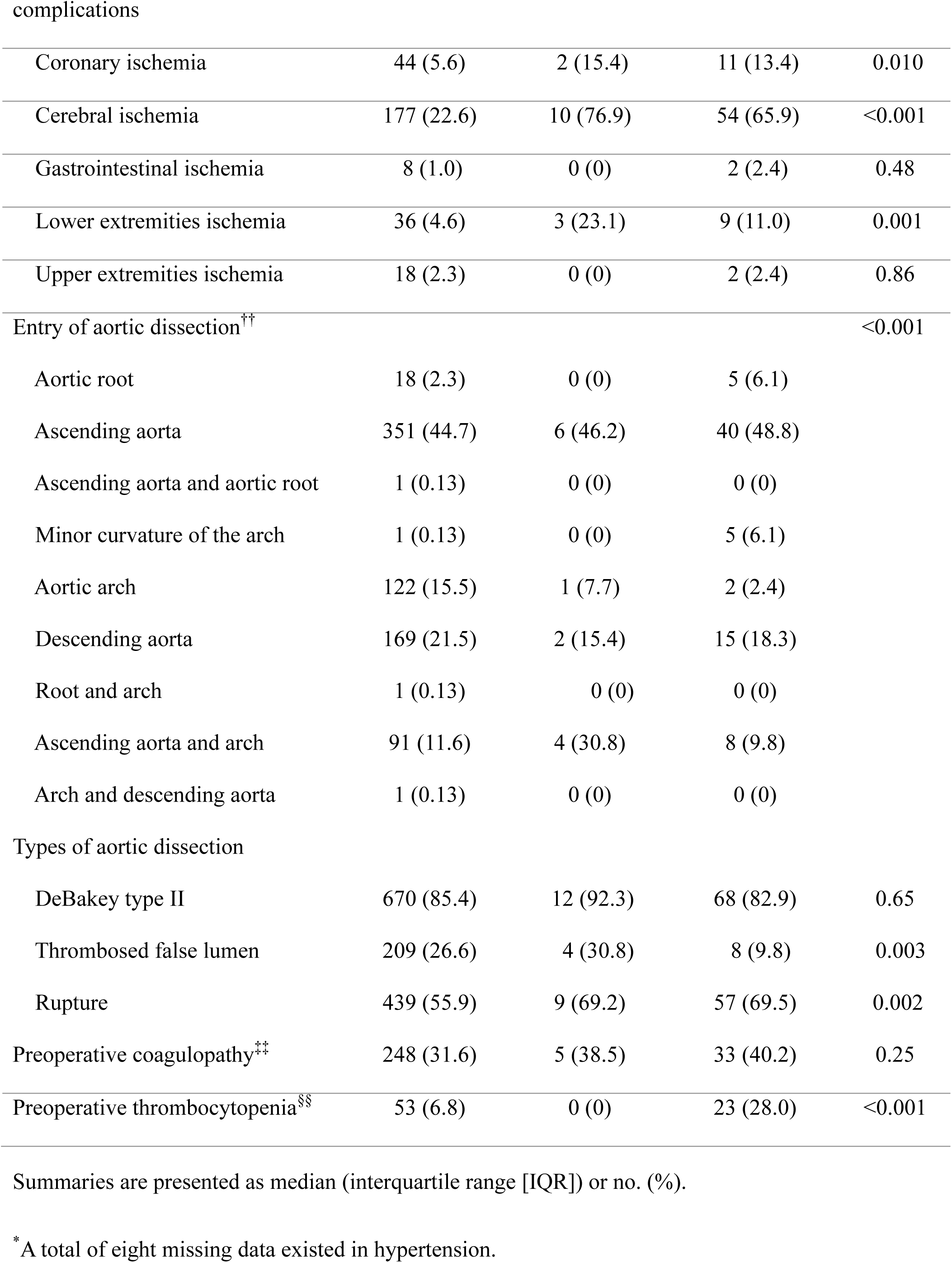
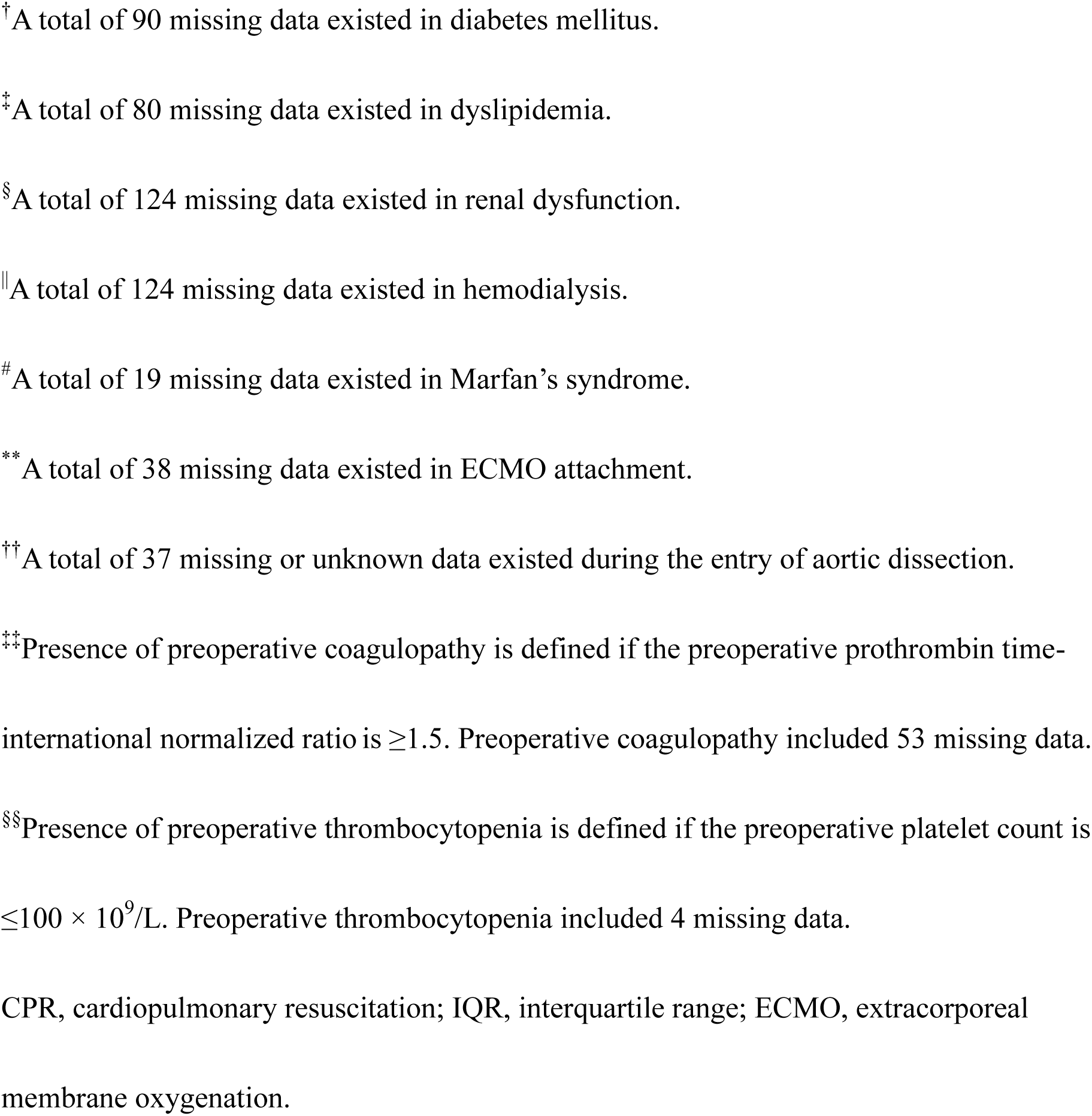
Patient characteristics of the three groups with CPR <15 or ≥15 min performed during preoperative or intraoperative periods (n=880)

**Table 2.**
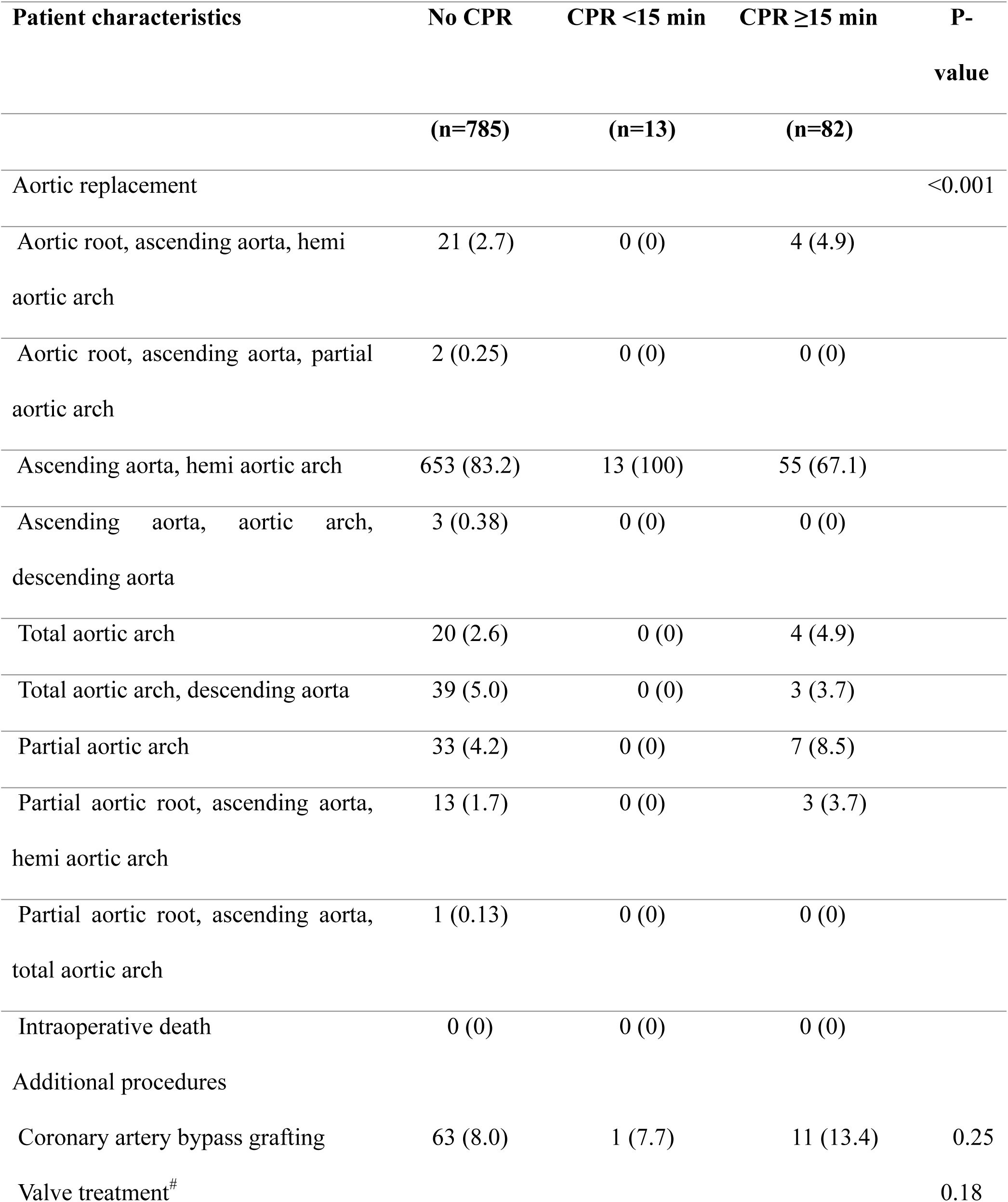
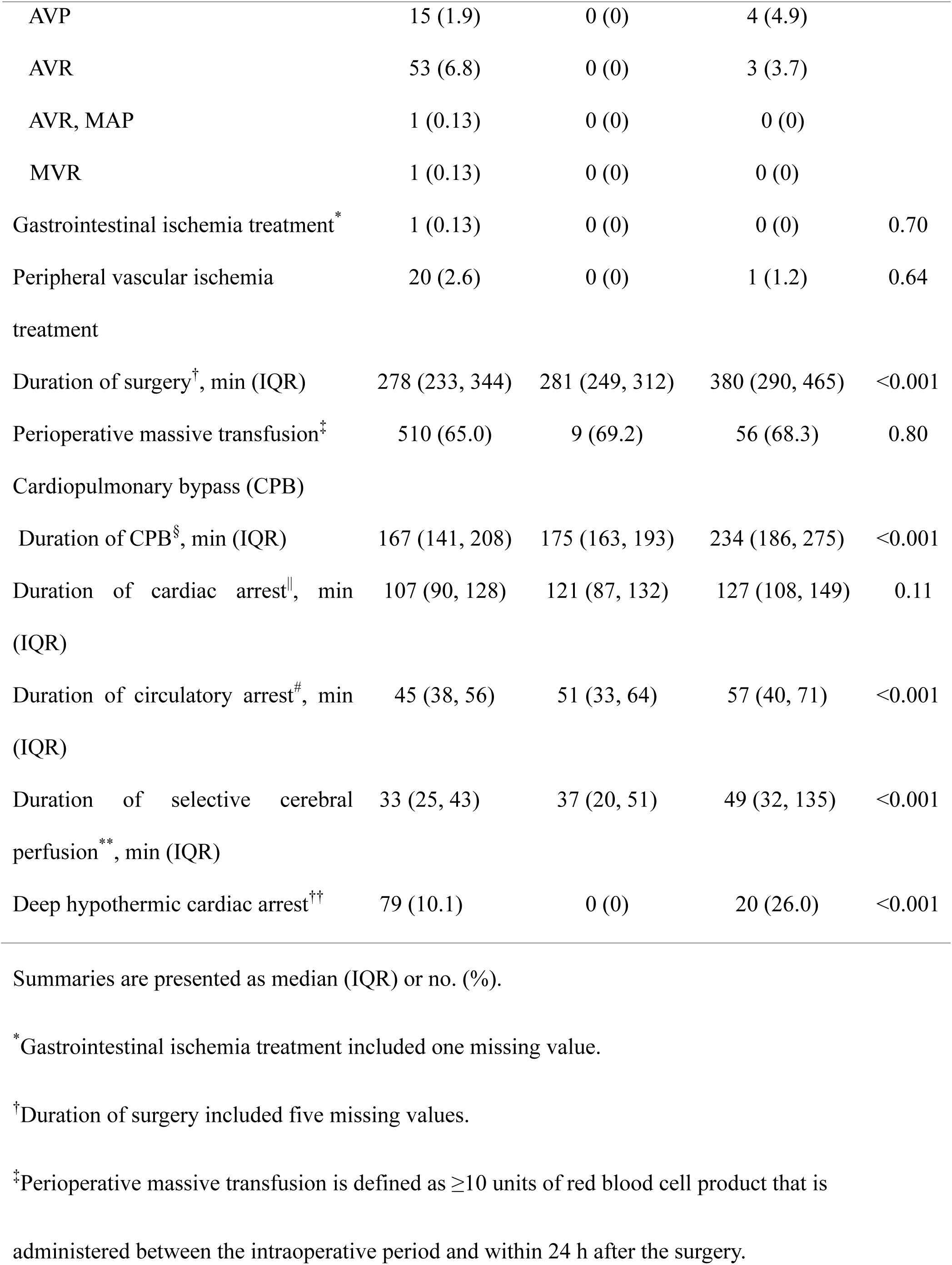
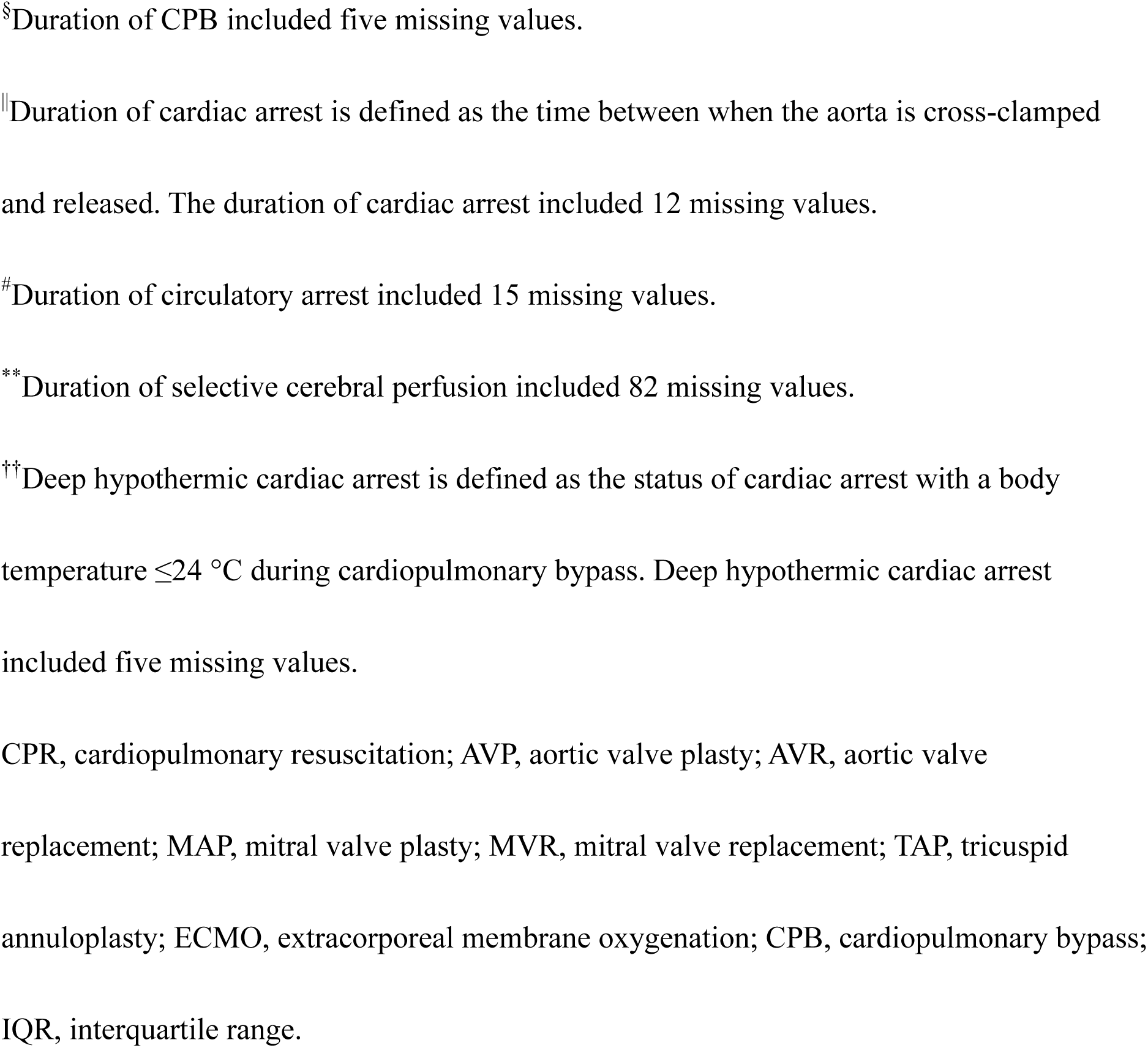
Surgical characteristics of the three groups with CPR <15 min and ≥15 min performed during preoperative or intraoperative periods (n=880)

**Table 3.** Postoperative complications in the three groups with CPR <15 min and ≥15 min performed during preoperative or intraoperative periods (n=880)

| Postoperative complications | No CPR<br>(n=785) | CPR <15 min<br>(n=13) | CPR ≥15 min<br>(n=82) | P-value |
| --- | --- | --- | --- | --- |
| Chest reopening | 64 (5.9) | 1 (7.7) | 3 (3.7) | <0.001 |
| Ventilatory support ≥1 week* | 86 (11.0) | 3 (23.1) | 12 (14.6) | <0.001 |
| Central nervous system disorders† |  |  |  |  |
| Focal neurological disorder | 67 (8.5) | 1 (7.7) | 6 (7.3) | <0.001 |
| Consciousness disorder | 48 (6.1) | 0 (0) | 2 (2.4) | <0.001 |
| Convulsion | 28 (3.6) | 0 (0) | 4 (4.9) | <0.001 |
| Others‡ | 38 (4.8) | 2 (15.4) | 6 (7.3) | <0.001 |
| Infections |  |  |  |  |
| Mediastinitis | 7 (0.89) | 0 (0) | 0 (0) | <0.001 |
| Surgical site infection | 48 (6.1) | 1 (7.7) | 3 (3.7) | <0.001 |
| Others§ | 26 (3.3) | 0 (0) | 2 (2.4) | <0.001 |
| Renal disorders |  |  |  | 0.001 |
| Renal failure without dialysis treatment | 319 (40.6) | 6 (46.2) | 48 (58.5) |  |
| Renal failure with<br>dialysis treatment | 25 (3.2) | 0 (0) | 7 (8.5) |  |
| Gastrointestinal<br>disorders | 11 (1.4) | 0 (0) | 1 (1.2) | 1.00 |
| Multiple organ<br>failure | 13 (1.7) | 0 (0) | 5 (6.1) | <0.001 |
| Extra circulation<br>support<sup>#</sup> | 20 (2.6) | 2 (15.4) | 16 (19.5) | <0.001 |
| Aortic complications<sup>**</sup> | 12 (1.5) | 0 (0) | 1 (1.2) | <0.001 |

Summaries are presented as no. (%).
\*Ventilatory support $\geq 1$ week included tracheal intubations and tracheostomy cases.
†Central nerve disorder counted cases without symptoms before surgery.
‡Other central nervous system disorders included cerebral infarction, dysphagia, cognitive dysfunction, visual field deficit, subarachnoid hemorrhage, cerebral hemorrhage, recurrent nerve paralysis, and neurogenic bladder.
§Other infections included sepsis, pneumonia, cholangitis, urinary tract infections, and vertebral osteomyelitis.
||Gastrointestinal disorders included gastrointestinal ischemic diseases, paralytic and obstructive ileus, hematochezia, and gastric ulcers.

### 3.2 Mortality within 30 days after Surgery

During the 10-year study period, death within 30 days after surgery occurred in 126/880 (14.3%). Death occurred in 76/785 (9.7%), 3/13 (23.1%), and 47/82 (57.3%) in patients without CPR events, with CPR <15 min, and ≥ 15 min, respectively.

Kaplan–Meier survival estimates within 30 days of surgery in the three groups with and without CPR events (none, CPR <15 min, and CPR ≥ 15 min) are shown in Figure 1. The survival functions were significantly different among the three groups (log-rank test: P<0.001). The unadjusted hazard ratios (HRs) of death within 30 days post-surgery were 2.82 (95% CI, 0.89–8.93; P=0.079) and 25.0 (95% CI, 17.0–36.8; P<0.001) in patients with CPR <15 min and CPR ≥15 min, respectively. Adjusted HRs (aHRs) and covariate-adjusted survival curves are shown in Table 4 and Figure 2. After adjusting for potential covariates, CPR administration for ≥15 min was associated with an increased risk of death within 30 days after surgery (aHR, 7.66; 95% CI, 3.56–16.5; P<0.001), but was not associated with CPR <15 min (aHR, 1.33; 95% CI, 0.37–4.78; P=0.66). However, as the group who received CPR <15 min was small, it could lead to non-significant results owing to insufficient power. There was an association between increased risk of death within 30 days after surgery and duration between preoperative ECMO attachment and CPB initiation ≥30 min, increase in age, presence of preoperative coronary ischemia and cardiac tamponade, preoperative renal dysfunction, and DeBakey type II dissection (Table 4). However, the presence of a thrombosed false lumen was associated with decreased risk of death within 30 days after surgery (Table 4).

**Figure 1.**
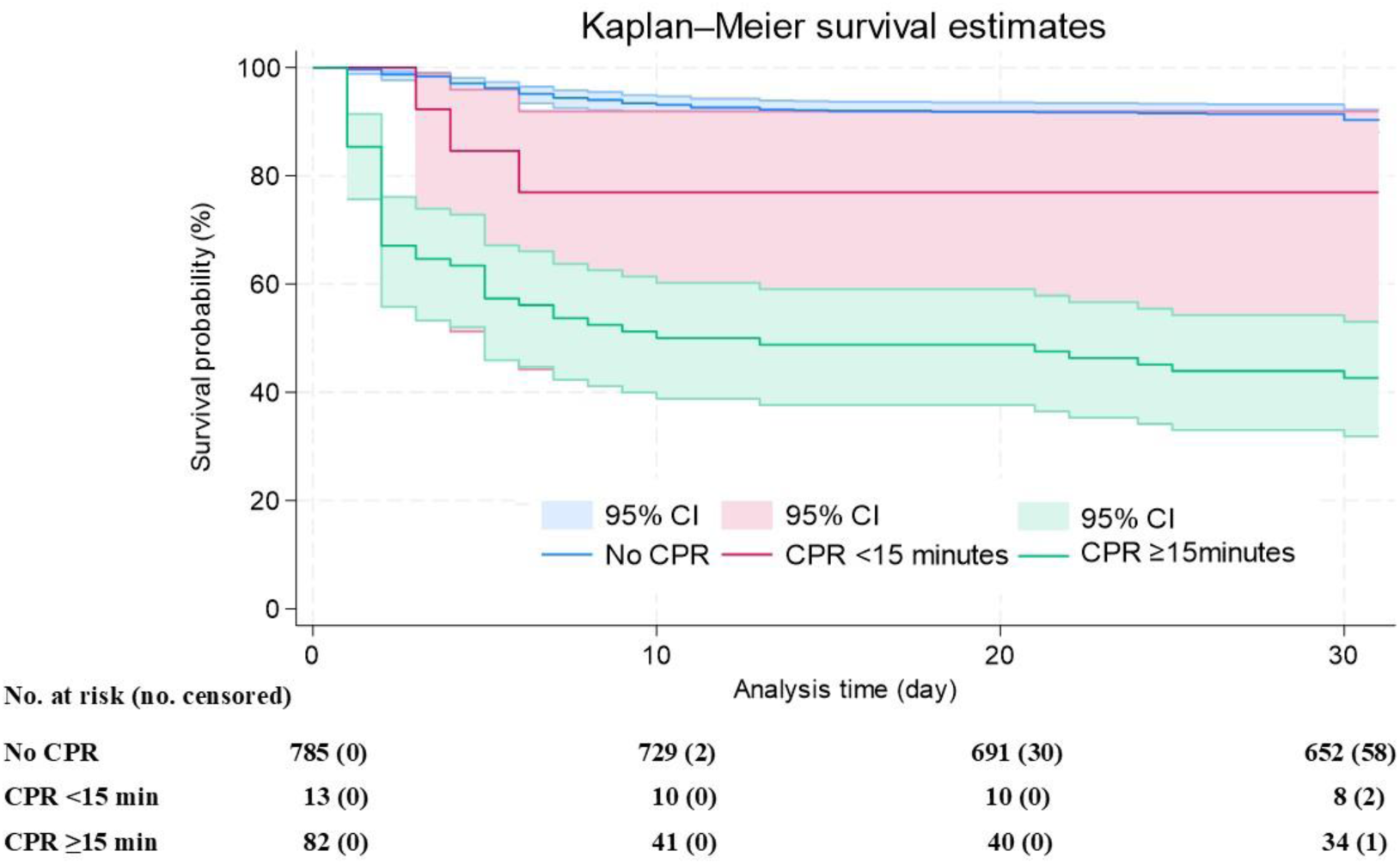
Unadjusted Kaplan–Meier survival estimates within 30 days postoperatively with 95% CI in the three CPR-duration groups (0, <15 min, or ≥15 min). The log-rank test shows a difference among the three groups (χ^2^=199.1; P<0.001). CI, confidence interval.

**Figure 2.**
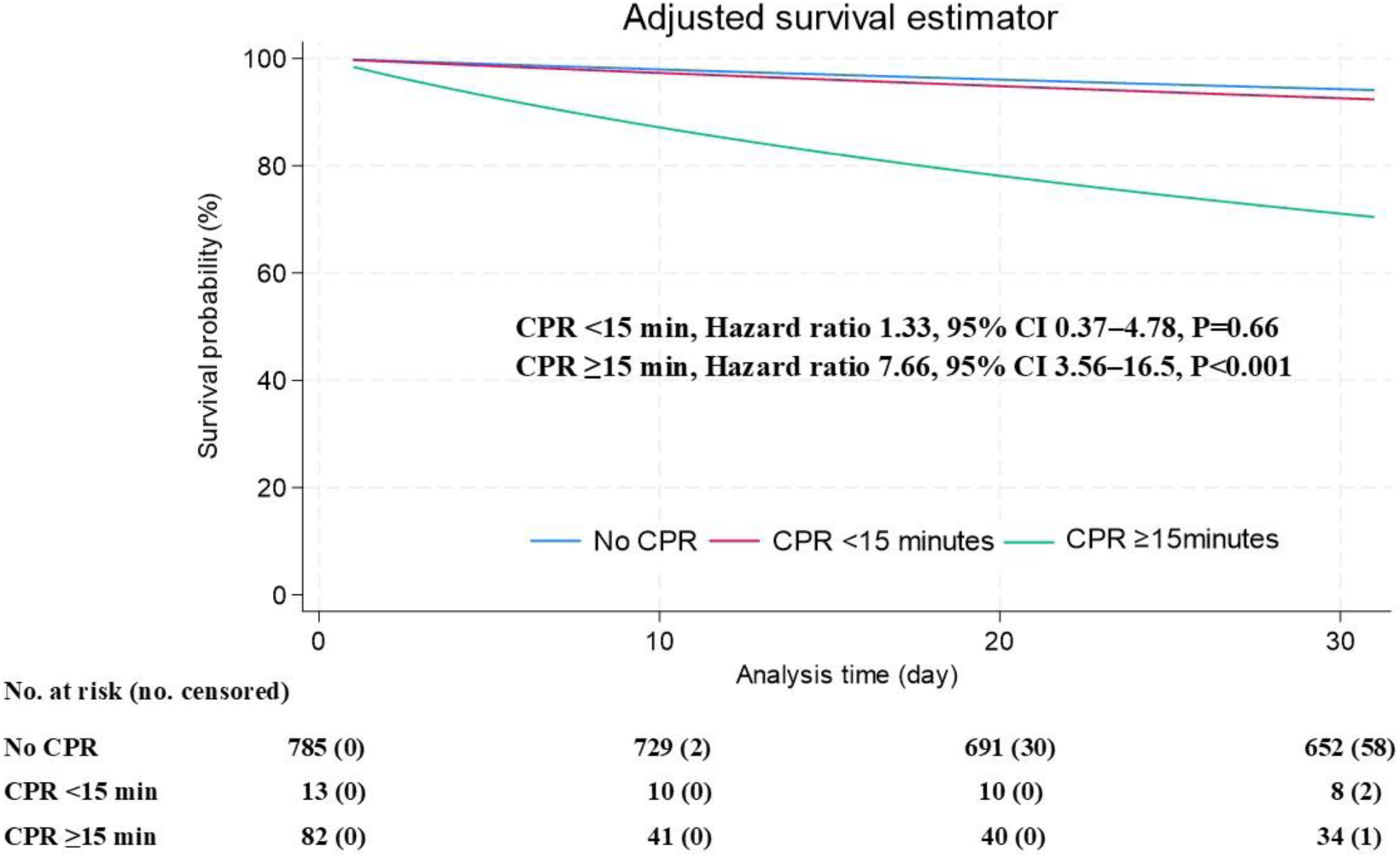
Adjusted survival estimator curves within 30 days postoperatively with a multilevel Cox hazards model based on the CPR-duration group. The probability is estimated with 95% CIs. CI, confidence interval.

**Table 4.**
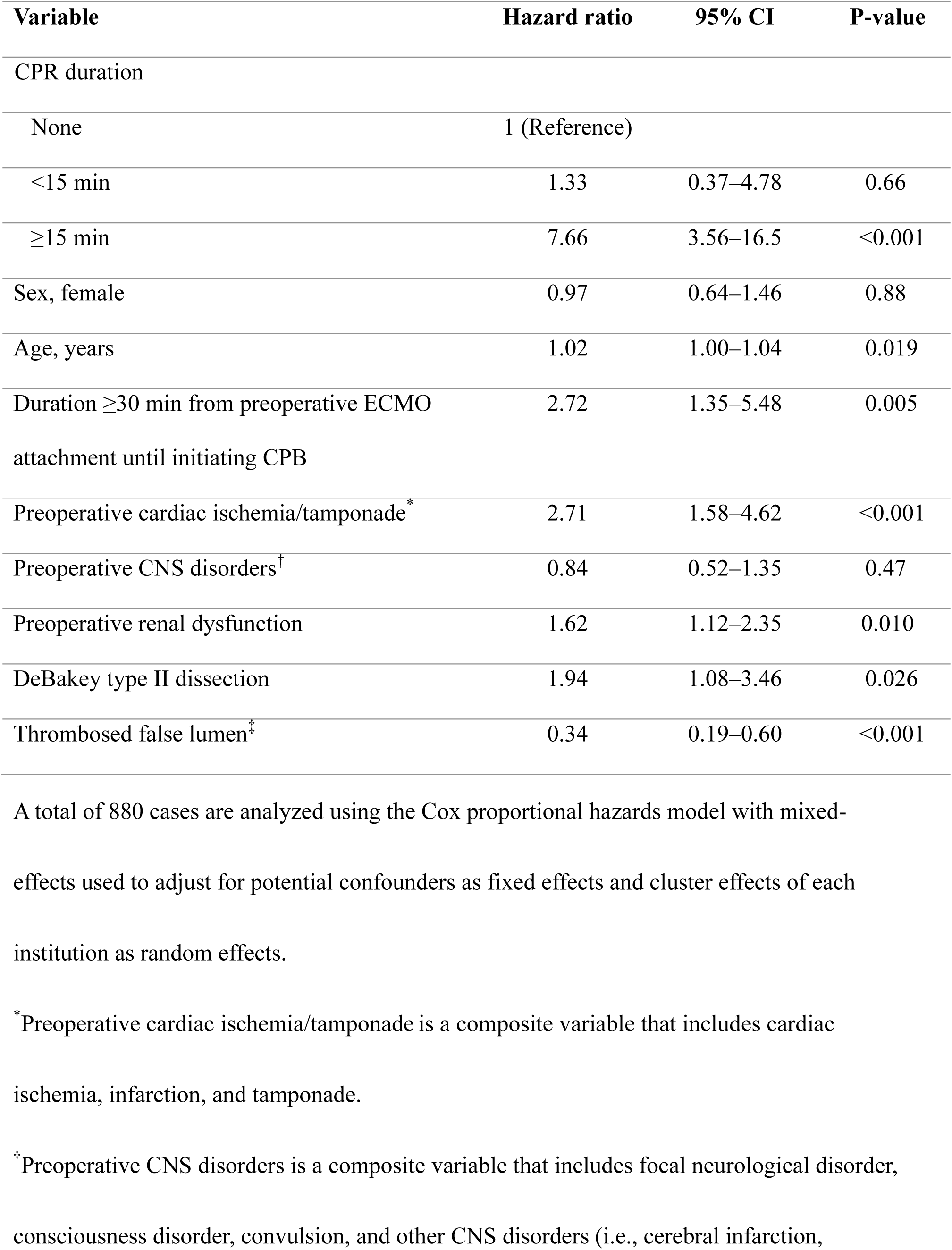
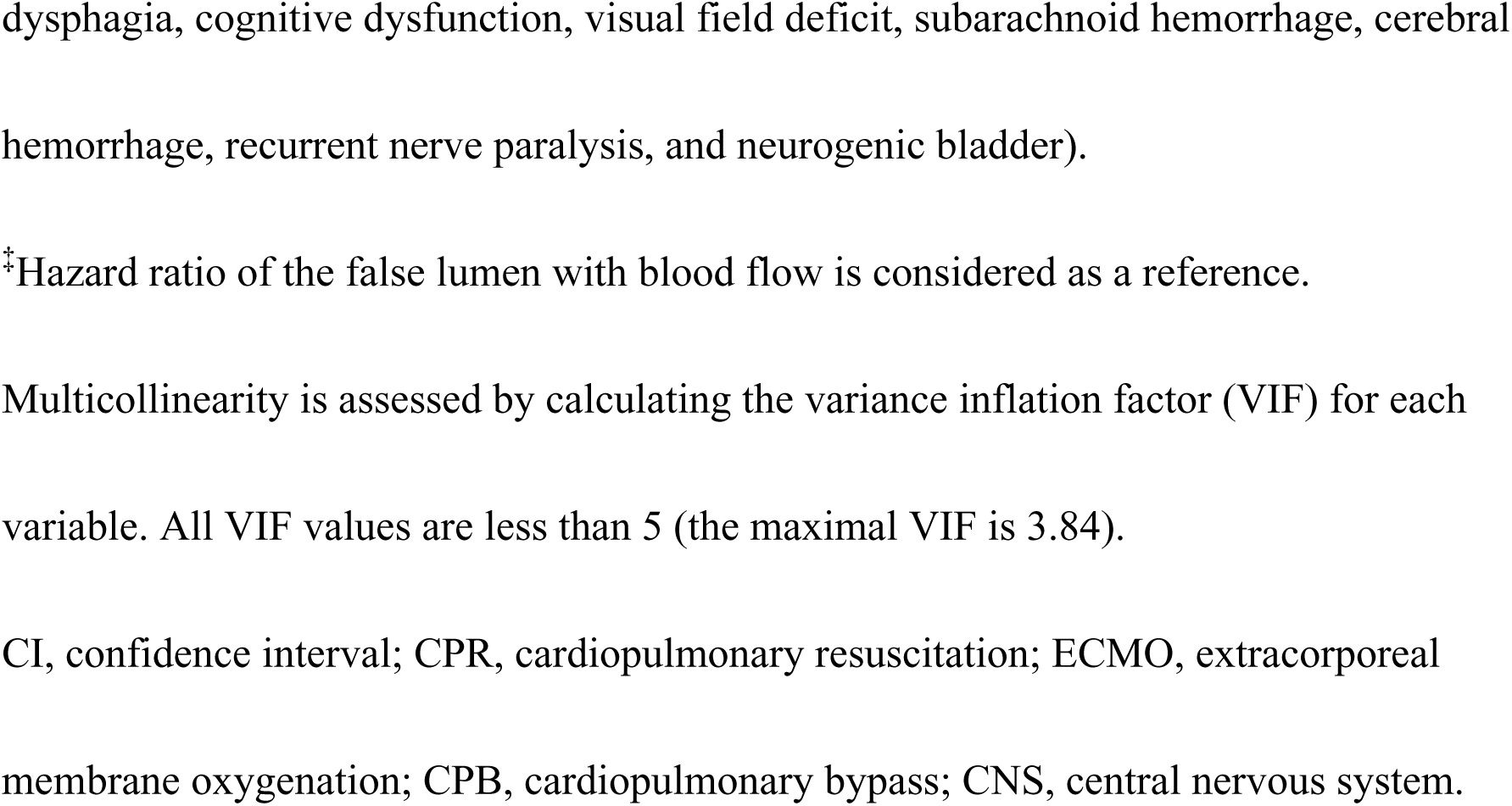
Hazard ratios for death within 30 days of surgery (n=880)

### 3.3 Secondary Outcomes

#### 3.3.1 Mortality within 7 days after Surgery

Death within 7 days after surgery occurred in 86/880 (9.8%). Death occurred in 45/785 (5.7%), 3/13 (23.1%), and 38/82 (46.3%) in patients without CPR events, with CPR <15 min, and ≥15 min, respectively.

The survival functions were significantly different among the three groups (log-rank test: P<0.001). The survival functions were significantly different among the three groups (log-rank test: P<0.001). The unadjusted HRs for death within 7 days post-surgery were 4.39 (95% CI, 1.36–14.1; P=0.013) and 22.8 (95% CI, 14.6–35.7; P<0.001) in patients with CPR <15 min and CPR ≥15 min, respectively. The aHRs and covariate-adjusted survival curves are shown in Table 5 and Figure 3. After adjusting for potential covariates, CPR administration for ≥15 min was associated with an increase in risk of death within 7 days after surgery (aHR, 6.29; 95% CI, 2.67–14.8; P<0.001), but not with CPR <15 min (aHR, 1.81; 95% CI, 0.49–6.76; P=0.38). There was an association between increased risk of death within 7 days after surgery and preoperative ECMO attachment and CPB initiation ≥30 min, presence of preoperative coronary ischemia and cardiac tamponade, and preoperative renal dysfunction (Table 5). However, the presence of a thrombosed false lumen was associated with decreased risk of death within 7 days after surgery (Table 5).

**Figure 3.**
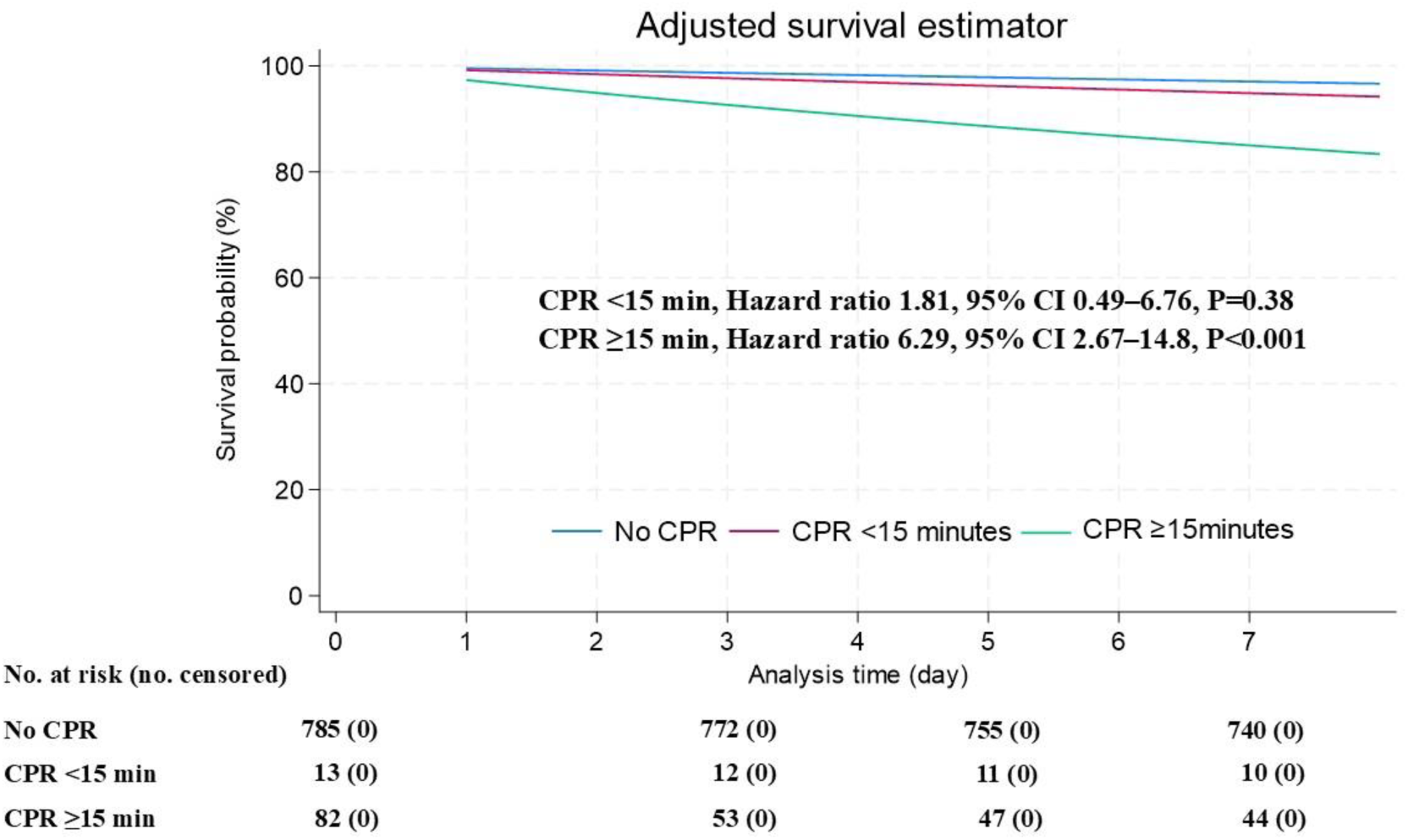
Adjusted survival estimator curves within 7 days postoperatively with a multilevel Cox hazards model based on the CPR-duration group. The probability is estimated with 95% CIs. CI, confidence interval.

**Table 5.**
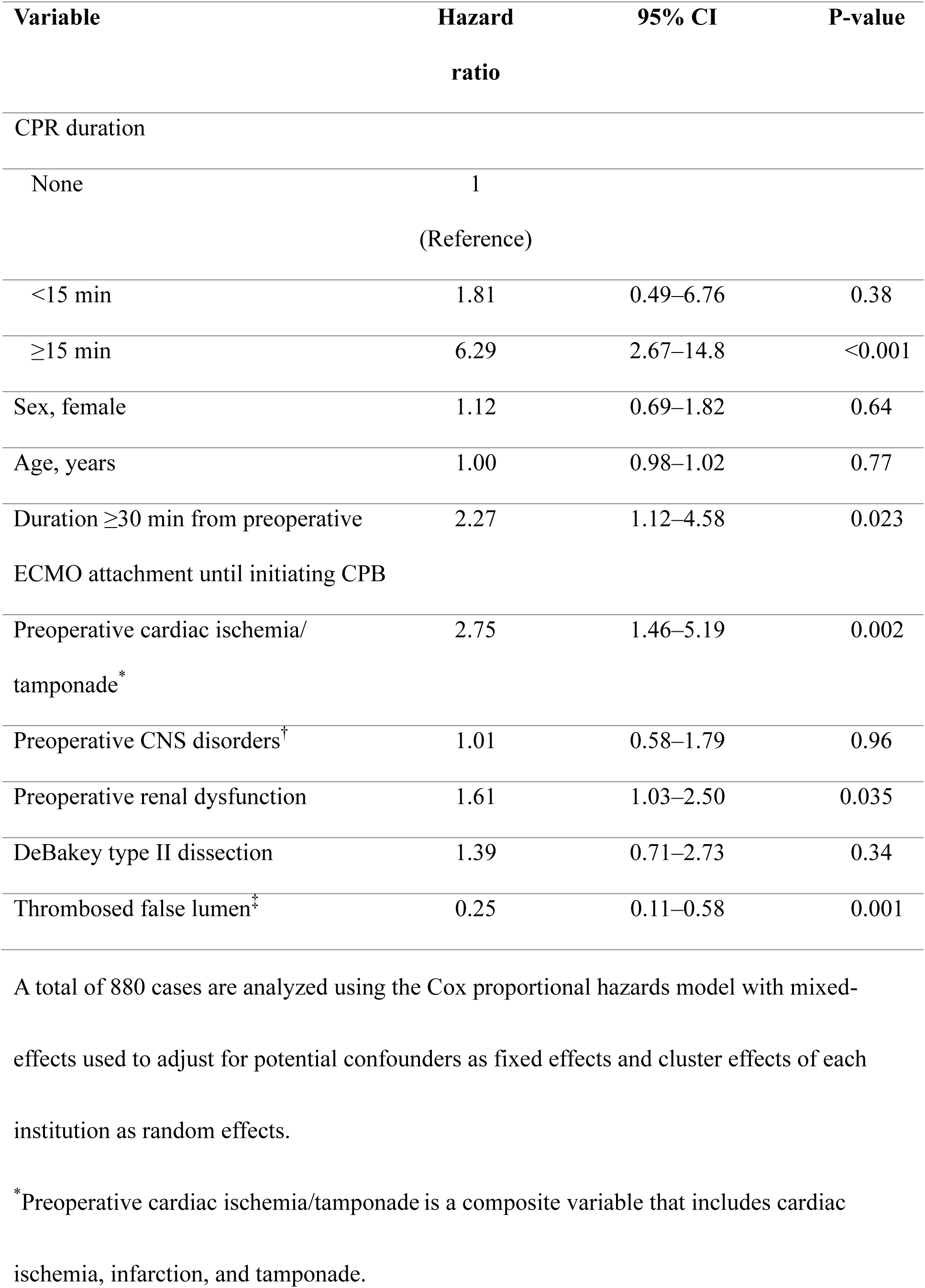
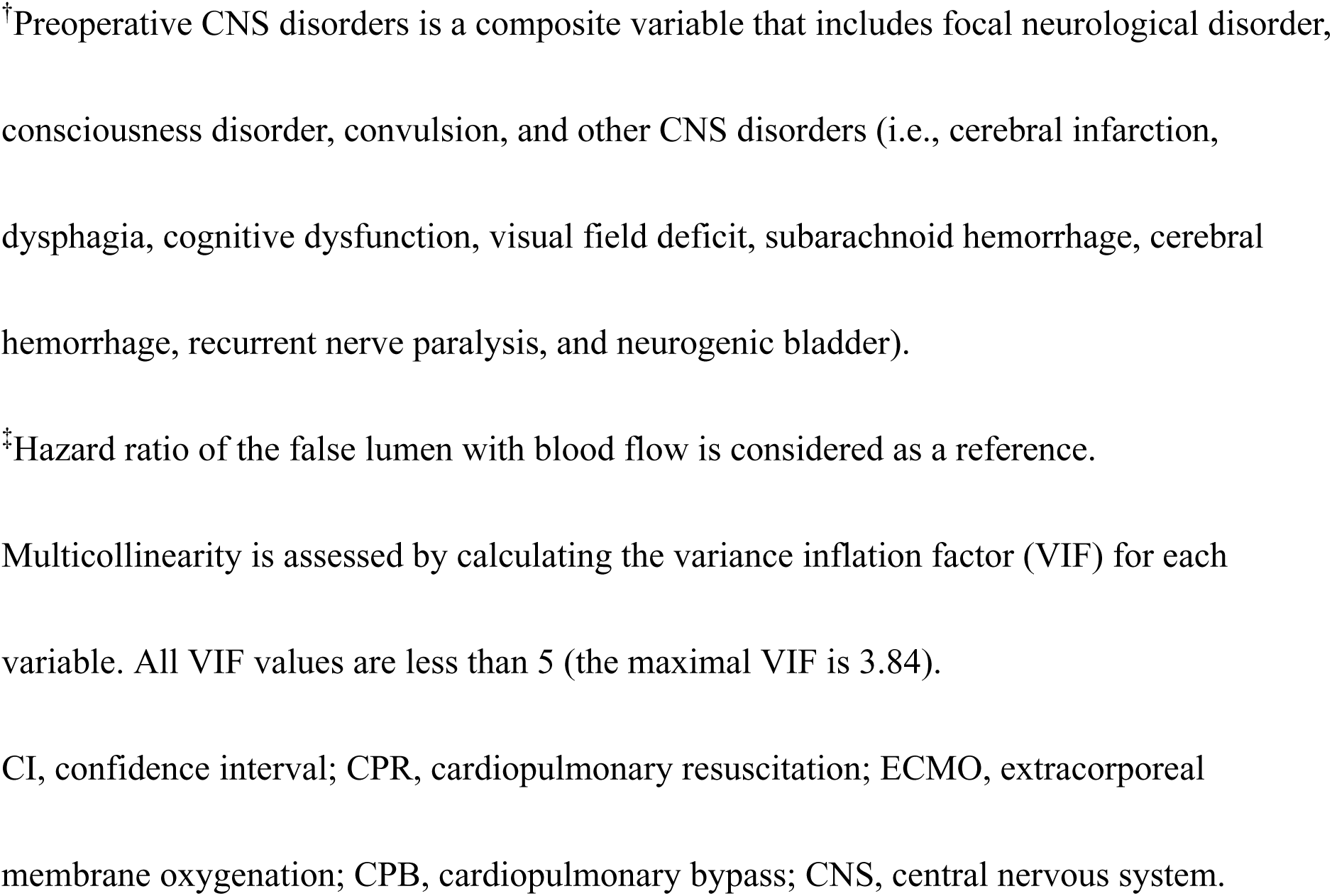
Hazard ratios for death within 7 days of surgery (n=880)

#### 3.3.2 CNS Complications within 30 days after Surgery

CNS complications within 30 days after surgery occurred in 184/880 (20.9%) patients. CNS complications occurred in 141/785 (18.0%), 5/13 (38.5%), and 38/82 (46.3%) in patients without CPR events, with CPR <15 min, and ≥15 min, respectively.

Competing-risks regression analysis showed that the unadjusted subhazard ratios (SHRs) of CNS complications within 30 days after surgery were 2.42 (95% CI, 2.42–2.43; P<0.001), and 1.36 (95% CI, 1.32–1.40; P<0.001) in patients with CPR <15 min and CPR ≥15 min, respectively. Adjusted SHRs (aSHRs) are shown in Supplemental Table S1. After adjusting for potential covariates, both CPR administration for <15 min and ≥15 min were associated with an increase in risk of CNS complications within 30 days after surgery (aSHR, 4.49; 95% CI, 3.92–5.11; P<0.001; aSHR, 3.62; 95% CI, 2.73–4.81; P<0.001, respectively). There was an association between increased risk of CNS complications within 30 days after surgery and an increase in age, preoperative renal dysfunction, and DeBakey type II dissection (Supplemental Table S1). However, there was an association between a decrease in the risk of CNS complications within 30 days after surgery and preoperative ECMO attachment and CPB initiation ≥30 min, female sex, presence of a thrombosed false lumen, and preoperative coronary ischemia and cardiac tamponade (Supplemental Table S1).

### 3.4 Sensitivity Analysis

The multilevel logistic regression analysis excluding participants with CPR <15 min showed that CPR administration for ≥15 min was consistently associated with an increase in risk of death within 30 days after surgery (aHR, 7.61; 95% CI, 3.57–16.2; P<0.001).

The sensitivity analysis excluding participants with ECMO attachment showed a consistent association between CPR administration for ≥15 min and 30-day postoperative mortality (aHR, 8.90; 95% CI, 4.20–18.8; P<0.001).

### 3.5 ECMO and CPR Duration in Patients with ECMO Attachment

The description regarding the duration of ECMO and CPR in patients with ECMO attachment are written in Supplemental Text 2.

## 4. DISCUSSION

This retrospective cohort study evaluated the association between CPR duration (< 15 or ≥15 min) and postoperative 30-day mortality at two Japanese tertiary-care hospitals. The study results showed that CPR performed for ≥15 min was associated with an increased risk of postoperative 30-day mortality. In addition, the postoperative 30-day mortality in patients with CPR lasting ≥15 min was approximately 480% higher than that in patients without CPR administration. Furthermore, despite its duration, CPR administration was associated with an increased risk of postoperative 30-day CNS complications.

There is no established consensus regarding the surgical indications for AAAD with CPA based on CPR duration. In Japan, the report from the Tokyo Acute Aortic Super-network indicates that 434/3,307 (13.1%) registered patients had CPA. The overall mortality of patients with AAAD along with CPA was 88.2% (94.5% of out-of-hospital CPA and 79.4% of in-hospital CPA).^5^ However, the acceptable duration of CPR has not been clearly defined. Shorter CPR duration has been correlated with higher survival rates.^22–24^ In the cohort of AAAD, previous reports from the Japanese national medical center also indicate that CPR duration exceeding 15 min was a significant risk for in-hospital mortality,^7^ and ROSC affected prognosis (median CPR time of 20 min).^8^ Several studies indicate that achieving ROSC before surgery directly correlates with improved survival and neurological outcomes.^6–9^ However, these studies were small-scale retrospective studies, and no established guidelines for the maximum duration of preoperative CPR or the limit of indication for AAAD with CPA are available. The present study suggested that a CPR duration of 15 min may represent a prognostic threshold. In our facility, to minimize CPR duration and increase survival, our top priority is to transport patients to the operating room as quickly as possible and promptly initiate CPB. To achieve this policy, it is essential to maintain close coordination with referring hospitals and establish standardized patient transport systems. Furthermore, we maintain full-time shifts of the emergency surgery team, including cardiac surgeons, anesthesiologists, and operating room staff, to respond to emergency surgeries.

Cardiac tamponade is the primary cause of CPA in AAAD. According to autopsy records in Japan, cardiac tamponade is the most common cause of death from AAAD for 85% of autopsy cases.^25^ In the present study, cardiac tamponade emerged as an independent predictor of increased 30-day postoperative mortality (HR=2.61), consistent with findings from the International Registry of Acute Aortic Dissection. Cardiac tamponade is a well-recognized marker of disease severity in AAAD and the principal contributor to early postoperative mortality.^26,27^ Under conditions of cardiac tamponade, the effectiveness of CPR is expected to be limited. In general, pericardial drainage is the standard treatment for cardiac tamponade.^28^ However, in AAAD, unregulated or rapid pericardial decompression may lead to abrupt increases in blood pressure, potentially promoting further bleeding or aortic re-rupture and adversely affecting outcomes.^29^ Reflecting these concerns, earlier European Society of Cardiology guidelines classified pericardial drainage in AAAD as a class III recommendation.^30^ Recently, however, several reports have suggested that a carefully titrated strategy—namely, controlled and intermittent pericardial drainage under strict blood pressure management—may be effective as a bridging measure to definitive surgical repair, with acceptable early and late outcomes in selected patients.^31,32^ At our institution, we prioritize immediate transfer to the operating room, where pericardial drainage can be performed under direct surgical control and prompt management for re-rupture is possible. Accordingly, pericardial drainage in the emergency department is generally avoided if it may delay definitive surgical repair. However, in selected cases—such as patients unresponsive to CPR or those who rapidly deteriorate into recurrent cardiac arrest after ROSC—limited pericardial drainage should be considered in the emergency department as a temporary measure.

The indication of ECMO attachment in patients with AAAD undergoing CPA is still controversial. Inconsistent results exist regarding ECMO’s protective effect on mortality and complications.^6,7,33,34^ ECMO is commonly applied to patients under CPA status without prior imaging studies, such as a contrast-enhanced CT scan. Consequently, attaching ECMO to patients with fragile aortic conditions can cause cerebral complications due to thrombus formation by the retrograde blood flow via the femoral artery. Furthermore, retrograde blood flow from the femoral artery may alter the pattern of aortic dissection, potentially causing new malperfusion or rupture.^6^ However, CPB, which is attached in the operating room, allows us to select a cannulation site without the femoral artery, such as the axillary artery, and to monitor changes in the aortic dissection, such as the form of false lumen or cerebral blood flow, via TEE or bibo-apical spectroscopy. Therefore, immediate CPB is preferable to ECMO initiation. Depending on the patient’s condition during transport and operating room availability, ECMO attachment can be a life-saving option. However, the usefulness of initiating ECMO in such situations remains unclear. Here, 14 patients required preoperative ECMO, of whom 12 (85.7%) died within 30 days, whereas two patients (14.3%) survived. Although the small sample size precludes formal statistical inference, several descriptive differences were observed between survivors and non-survivors. The duration of ECMO support was approximately four-fold longer in the survivor group, and the interval from initiation of CPR to ECMO deployment was approximately half of that observed in non-survivors. These observations may suggest that earlier initiation of ECMO is associated with improved survival, even when the interval to surgery is prolonged in patients who do not achieve prompt ROSC or when definitive surgical repair is unavoidably delayed.

Nevertheless, these findings should be interpreted with caution. Although ECMO support is expected to improve perfusion to vital organs compared with conventional CPR, the present study does not allow definitive conclusions regarding a survival benefit associated with ECMO use. Furthermore, ECMO support and ROSC represent fundamentally different physiological states, and outcomes under ECMO cannot be considered equivalent to those achieved after ROSC.

Based on the sensitivity analyses, the principal finding of this study is that a CPR duration of 15 min or longer is associated with an increased risk of mortality. Further investigations are warranted to clarify the extent to which ECMO support modifies this association.

### 4.1 Limitations

Despite the present study providing clinically relevant insights into one of the most challenging scenarios in aortic surgery and highlighting critical areas for future investigation, it has some important limitations. First, the exposure variable was defined as the combined duration of pre- and intra-operative CPR time. This requires careful interpretation, since CPR administration can be provided in different situations in prehospital and operative settings.

Two clinically important time points guide whether to proceed with surgery: prior to initiation of surgery and before CPB initiation. Thus, our definition appears clinically optimal, but requires cautious interpretation owing to its heterogeneity. Second, the sample size of the group with CPR duration <15 min was small, which could induce instability in regression models and inflate type II errors. By conducting a sensitivity analysis excluding the cohort with CPR time <15 min, consistent results were obtained. However, the risk associated with CPR time <15 min for postoperative mortality was inconclusive. Third, by design, the policy of discontinuing transfer after extended CPR attempts biases the study cohort toward survivors, thereby yielding results toward better outcomes through systematic selection bias. As a result, our findings may be biased toward the null, and the actual risk could be higher than that reported. Further, the association for CPR duration <15 min was inconclusive in this study, and may have been biased toward the null owing to survival bias. Fourth, the results are subject to bias from unmeasured confounding despite careful data collection and analysis. Fifth, our findings are limited in generalizability because the data were obtained from a limited number of Japanese institutions. However, our findings can provide essential information regarding the indication of surgery in patients with AAAD undergoing CPA. Sixth, misclassification bias can distort our results. In the present study, all patients with acute aortic dissection complicated by CPA did not necessarily receive an accurate diagnosis before death. Some patients died prior to hospital arrival or due to the absence of definitive imaging confirmation. Finally, perioperative management strategies—particularly the use of ECMO and other advanced resuscitative measures—were not standardized; they were determined on a case-by-case basis at the treating team’s discretion. Because standardized criteria or a uniform treatment algorithm were not applied, variability in clinical decision-making may have influenced outcomes. This heterogeneity limits the ability to draw definitive conclusions regarding the efficacy or optimal timing of ECMO and underscores the need for prospective studies with predefined management strategies.

### 4.2 Conclusion

In AAAD with preoperative CPA, CPR duration ≥15 min was associated with increased risk of postoperative 30-day mortality. In addition, CPR performance despite the duration was associated with early postoperative CNS complications. Surgical indications for patients with CPR ≥15 min must be carefully discussed. Although a preoperative CPR duration of 15 min may represent a potential threshold for surgical decision-making, outcomes among patients with CPR duration of less than 15 min remain uncertain, and no definitive conclusions can be drawn from the present study. Further investigation into the impact of early ECMO attachment for reducing CPR duration on postoperative mortality is desired.

## Non-standard Abbreviations and Acronyms

aHR: adjusted hazard ratio
CI: confidence interval
CNS: central nervous system
CPAOA: cardiopulmonary arrest on arrival
CPB: cardiopulmonary bypass
CPR: cardiopulmonary resuscitation
CT: computed tomography
ECOM: extracorporeal membrane oxygenation
HR: hazard ratio
IQR: interquartile range
ROSC: return of spontaneous circulation

## Acknowledgements

We would like to thank Editage (www.editage.jp) for the professional English language editing service.

## Data availability statement

The data are not publicly available because of ethical and legal restrictions. De-identified data supporting the findings of this study are available from the corresponding author upon reasonable requests.

## Supplemental Material

Supplemental Text 1–2

Table S1 References 35–36

All of these supplemental materials are for online publication.

